# Age distribution characteristics of 1915 cases of otitis media with effusion in children

**DOI:** 10.1101/2024.09.03.24313029

**Authors:** Mingyang Li, Xiangyu Chen, Jingyue Xu, Zihan Liu, Cuncun Xie, Xiaodong Jia, Shaoguang Ding, Guangke Wang, Hongjian Liu

## Abstract

**Objective:** Objective To study the age distribution characteristics of otitis media with effusion in children and provide a basis for the selection of the timing of tympanic membrane perforation repair in children.

**Methods:** A total of 1915 children with otitis media with effusion who visited our hospital from 2017 to 2022 were collected and divided into 4 groups according to age, namely, infant group, preschool group, school-age group and adolescent group, to study the distribution characteristics of the age of onset of children with otitis media with effusion.

**Results:** The peak age of children with otitis media with effusion in the preschool period of 4-6 years old was the peak age of children with otitis media with effusion. The number of visits for otitis media with effusion in children aged 7 years old showed an obvious decline “inflection point”, and the number of visits for otitis media with effusion in adolescent children after the age of 13 years old tended to be stable.

**Conclusion:** The incidence of otitis media with effusion in children decreased significantly after the age of 7, and the number of visits for otitis media with effusion in children aged 13 years was close to that of adults. This conclusion provides a theoretical basis for the timing of tympanic membrane perforation repair in children.

**Short summary:** This study aims to investigate the characteristics of otitis media with effusion otitis media with effusion (OME) in children, focusing on the peak periods for treatment and surgical intervention, the “inflection point” in treatment trends, and the overall changing trends in OME. The findings will provide a foundation for determining the appropriate timing for surgical intervention in pediatric OME cases.

## Level of Evidence: 4

As endoscopic ear surgery advances, the age at which children undergo tympanoplasty is decreasing; however, the optimal timing for this procedure remains a subject of debate. A study involving 149 patients aged 5-17 years (mean age of 10.5 years) who underwent tympanoplasty reported a 91.3% success rate at six-month follow-up, with a recurrence rate of 35.3% in patients under 7 years of age ^[1]^. Additional studies indicate that the success rate of tympanoplasty in children under 4 years of age is below 26% ^[2]^. The leading cause of tympanoplasty failure in children is re-perforation due to middle ear infections, with a significant proportion attributed to OME ^[3, 4]^. Currently, the majority of OME patients are children. Research has estimated the incidence rate among 2-year-olds in Europe and the United States at 3.8% ^[3]^; however, accurate statistical data on the age of onset for Asian children remain scarce. Understanding the age-related characteristics of OME in children is crucial for determining the appropriate timing for tympanoplasty. This study collected and analyzed 1915 pediatric cases of OME from a clinical epidemiological perspective, focusing on the age distribution characteristics.

## 1 Materials and Methods

*Name of ethics committee:Medical Ethics Committee of HenanProvincial People’s Hospital. Approval number: 2019PHB109-01*.

### 1.1 Inclusion criteria for patients with otitis media with effusion

According to the diagnostic criteria for otitis media with effusion ^[5,6]^, 1915 patients aged 1 to 18 years who visited our hospital for otitis media with effusion from 2017 to 2022 were collected. All 1915 patients met the following conditions: 1. Otoscopic examination showed fluid accumulation in the middle ear 2. The patient’s acoustic impedance test showed a “B” or “C” curve, and children under 6 months old had no positive peak in the 1000 Hz probe tone test 3. There was a bone-air difference in behavioral audiometry or auditory brainstem response (ABR) bone-air threshold test.

### 1.2 Patient grouping

In accordance with the Law of the People’s Republic of China on the Protection of Minors^[7]^, the United Nations Convention on the Rights of the Child^[8]^, and various authoritative international and domestic sources, children are defined as individuals under the age of 18. According to the ninth edition of the People’s Health Education Press textbook Pediatrics and the World Health Organization, children aged 1-3 years are classified as infants, 4-6 years as preschoolers, 7-13 years as school-age children, and 14-18 years as adolescents^[9,10]^. Data on the number of children undergoing surgery in outpatient clinics and wards across each age group were collected in integer age units, and changes in data between different age groups were analyzed.

### 1.3 Study on the “turning point” of the decline in the age of children seeking medical treatment

The number of children in each age group was counted separately, with data summarized according to the number of children in each age category. The slope *k* for each point was calculated using the formula *k*=*y*2−*y*1/*x*2−*x*1, where *y*2−*y*1represents the difference in the number of children at adjacent ages, and *x*2−*x*1represents the difference in age. The point with the largest negative slope was identified as the “inflection point” ^[11]^, indicating the age at which the decline in the number of children with otitis media with effusion is most pronounced.

### 1.4 Statistical Methods

Statistical analyses were performed using SPSS software. Quantitative data conforming to a normal distribution were described using the mean ± standard deviation, skewed quantitative data were reported as median ± quartile, and qualitative data were presented as composition ratios or percentages. Single-factor ANOVA was employed to analyze data across different age groups, with multiple comparisons conducted to determine statistical differences between the groups. A p-value of less than 0.05 was considered statistically significant for all analyses.

## 2 Results

### 2.1 Conditions of children in each age group

There were 1915 children with otitis media with effusion. According to the age groups, there were 247 children aged 1-3 years, 797 preschoolers aged 4-6 years,707 schoolchildren aged 7-13 years, and 164 adolescents aged 14-18 years, as shown in Table 1. The overall trend of otitis media with effusion in children was first rising and then falling. The age group with the most cases was 4-6 years old in the preschool group. The incidence of otitis media with effusion in schoolchildren and adolescents gradually decreased and stabilized with age. There was a significant statistical difference between preschool children and the other three groups in the comparison of age groups (P < 0.001).

**Table 1.** Primary data of children of different ages groups.

|  | Age Group | Number of procedures | Number of patient X±s | P |
| --- | --- | --- | --- | --- |
| Toddler | 1-3 Y | 247 | 82.33±62.164 | P<0.001 |
| Preschool children * | 4-6 Y | 797 | 265.66±19.655 |  |
| School-age children | 7-13 Y | 707 | 101.00±38.738 |  |
| Adolescent children | 14-18 Y | 164 | 32.80±8.526 |  |

\*Statistical significance at $P < 0.05$ .
X±s Mean plus or minus standard deviation.

### 2.2 Analysis of the “turning point” of patients’ age at consultation

This study utilized slope analysis to estimate the trends in the number of children with otitis media with effusion across different ages. The point with the steepest negative slope was identified as the “inflection point,” indicating the most significant decline at age 7, The data can be seen in Figure 1 below.

**Figure 1.**
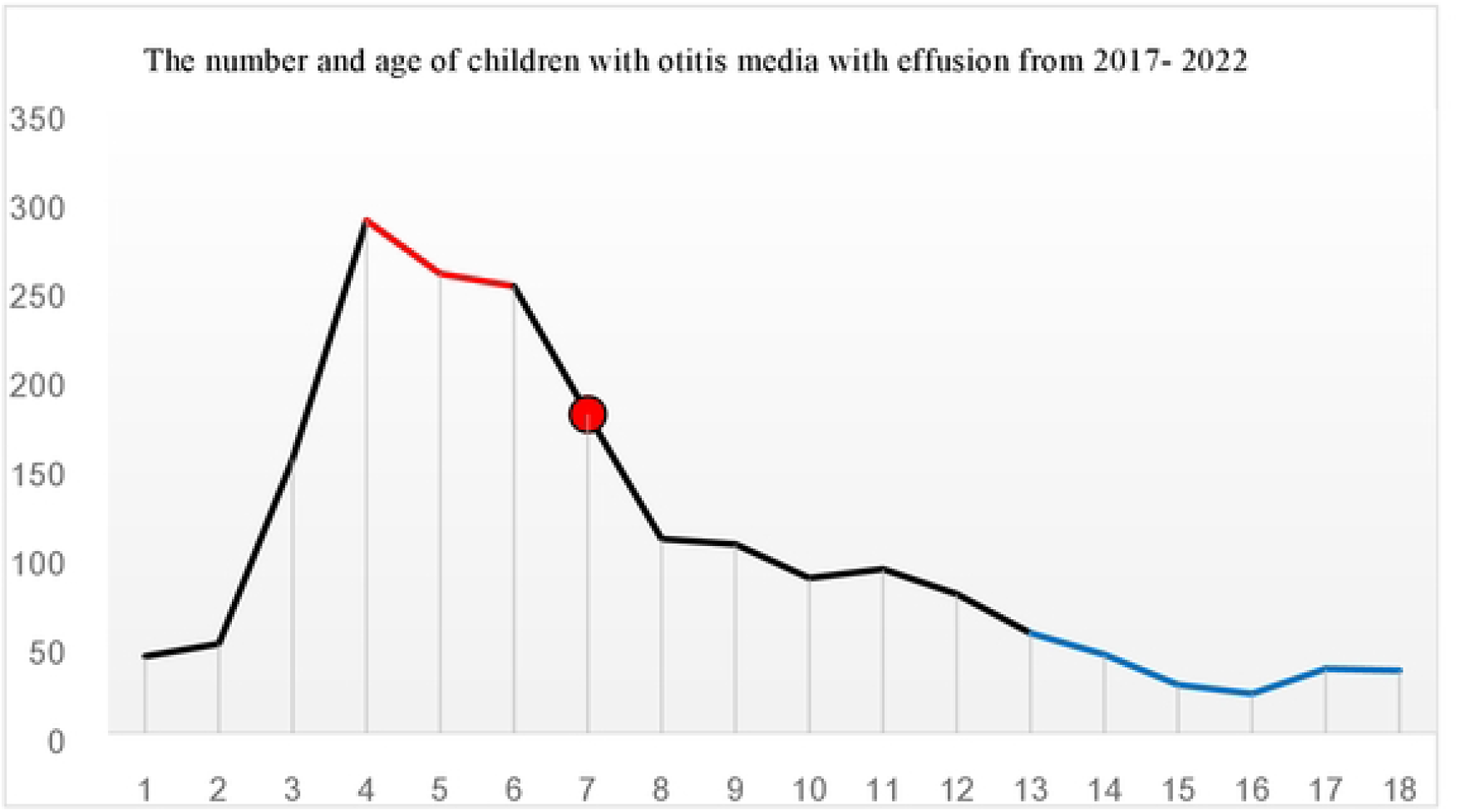
Changes in the numberof ehi.ldren with otitis media witheffusion in 2017 – 2022

## 3 Discussion

The incidence of otitis media with effusion (OME) in children in Europe and the United States is currently reported to be between 8.7% and 18.3%^[5]^; however, comparable statistics for Asian countries are lacking.

Otitis media with effusion (OME) is a known cause of hearing loss in children. In this study, the incidence of OME among children aged 1-2 years was relatively low. This may be attributed to the immune protective factors acquired through breastfeeding, which enhance individual immunity and reduce the incidence of respiratory tract infections ^[12-14]^, including otitis media ^[15,16]^. OMEe scholars have suggested that the high-incidence age for chronic OME is between 5 and 7 years, with peak onset periods at ages 2 and 5 ^[17]^. In this study, the highest number of OME cases was observed in children aged 4-6 years, identifying preschool children as the high-incidence age group. Age 7 represents the “turning point” at which the number of OME visits begins to decline. As children age, the incidence of OME gradually decreases. Most clinical studies attribute the high incidence of OME in children to the immature development, incomplete function, and dysfunction of the Eustachian tube. The shorter and straighter Eustachian tubes in children, compared to adults, make them more susceptible to gastric acid reflux and subsequent inflammation, increasing the risk of otitis media^[18]^. The Eustachian tube typically reaches adult levels of development around age 7^[2]^.

This study found that the number of children with otitis media with effusion tends to stabilize after the age of 13. OMEe scholars believe that the high incidence of otitis media with effusion in children is mainly due to hypertrophy of the adenoids. Hypertrophic adenoids are prone to block the Eustachian tube and are more likely to retain microorganisms around the adenoids, i.e., near the Eustachian tube, further leading to otitis media with effusion ^[19]^. Adenoids generally begin to degenerate at the age of 8 and do not reach a completely stable state until the age of 13. Mishra et al. compared the middle ear pressure of children and adults at different ages and found that the middle ear pressure of children was close to that of adults at the age of 10-12 ^[20]^.

There are many reasons for the low success rate of surgical repair of tympanic membrane perforation in children. Most scholars believe that postoperative otitis media causes re-perforation of the tympanic membrane. The main cause of otitis media in children is the physiological development of the Eustachian tube. The function of the Eustachian tube reaches the adult level at the age of 7-8 years. Therefore, tympanoplasty should be performed with caution in children under the age of seven. OMEe scholars have proposed that otitis media surgery is not recommended for preschool children before the age of 6 years ^[2]^. This study found that the number of visits for otitis media with effusion in children showed an “inflection point” at the age of 7 years. Therefore, it is recommended that 7 years old be used as the dividing age for conservative and surgical treatment. That is, conservative treatment can be adopted before the age of 7 ^[21]^; surgical treatment can be adopted after the age of 7 years. herefore, it is recommended to perform tympanoplasty after the age of 7 years to reduce the incidence of postoperative otitis media with effusion. In addition, according to child development psychology, surgical trauma should be avoided as much as possible for children between the ages of 3 and 7 years. The mental development of preschool children is not mature yet, and their understanding of the things around them is not mature. Surgery at this age will increase the uncertainty of surgical prognosis ^[22]^. The appropriate timing of surgery has an important impact on the physical and mental growth of children.

## 4 Conclusion

The study results indicate that the peak incidence of otitis media with effusion (OME) occurs in children aged 4-6 years, with age 7 serving as the “turning point” for a decline in OME cases, and a stabilization in the number of cases observed at age 13. Based on these findings, it is recommended that age 7 be considered as a key determinant in deciding the timing of tympanic membrane perforation surgery in children.

## Data Availability

All relevant data are within the manuscript and its Supporting Information files.

